# COVID specificity of air pollution associations with mortality in Cook County Medical Examiner records

**DOI:** 10.64898/2026.11.24.26368859

**Authors:** Adam Florzak, Jonathan H. Tomkin

**Affiliations:** School of Earth, Society and Environment, University of Illinois Urbana-Champaign, Urbana, Illinois, USA

**Keywords:** COVID-19, fine particulate matter, nitrogen dioxide, exposure assessment, spatial confounding, Monte Carlo simulation

## Abstract

**Background:** The double-hit hypothesis proposes that chronic fine particulate matter (PM_2.5_) exposure and acute nitrogen dioxide (NO_2_) exposure increase COVID-19 mortality. Spatially patterned pollution and mortality complicate interpretation of these associations.

**Objective:** To test whether pollution exposure was more strongly associated with COVID deaths than with other natural deaths in Cook County Medical Examiner records.

**Methods:** We linked death records with demographic characteristics, Census population data, and modeled and monitored pollution estimates. Adjusted logistic regression compared 10,887 COVID deaths with 4,123 contemporaneous non-COVID natural deaths during March 2020–August 2021; sensitivity analysis used 12,048 pre-pandemic non-COVID deaths. Population-weighted Monte Carlo analyses compared observed exposures with group-specific expectations. Separate spatial and temporal randomizations examined short-term NO_2_ exposure.

**Results:** Higher five-year PM_2.5_ was associated with lower odds that a death record was classified as COVID rather than non-COVID (odds ratio per 1 µg/m³, 0.63; 95% confidence interval, 0.55–0.72), with a similar pre-pandemic comparison (0.54; 0.48–0.60). Modeled PM_2.5_ exceeded population-weighted expectations in both death groups. Modeled short-term NO_2_ excesses were small for COVID deaths and consistently larger for contemporaneous non-COVID deaths. At actual death times, NO_2_ exposure at death locations exceeded the population-weighted spatial expectation. At actual death locations, exposure immediately before death did not exceed the expectation based on randomized nearby times.

**Significance:** Pollution associations in these records were not specific to COVID deaths. The NO_2_ comparisons supported a spatial interpretation without evidence of unusually elevated exposure immediately before death. The study does not estimate population-level mortality risk.

## Introduction

Frontera et al. [1] described this as the “double-hit” hypothesis: long-term exposure to particulate aerosols followed by acute exposure to nitrogen dioxide. Ecological studies of long-term PM_2.5_ and NO_2_ exposure provide related evidence for its component pollution claims [2, 3].

In this model, persons chronically exposed to high PM_2.5_ concentrations overexpress angiotensin-converting enzyme 2 (ACE2), and this facilitates COVID-19 viral penetration. Experimental work in mice subsequently found that exposure to particulate matter increased pulmonary expression of ACE2 and TMPRSS2, providing biological plausibility for this pathway [4]. The reduction in normal ACE2 activity resulting from COVID-19 infection could then result in clinical deterioration by disrupting dependent cardiometabolic pathways. Once bound to ACE2, COVID-19 downregulates cellular expression of ACE2, also resulting in dysregulated Ang-II and proinflammatory lung response [5]. Once infected, the second hit comes from high-end episodic exposure to NO_2_, which causes an inflammatory response and oxidative stress that may induce respiratory distress [1].

The “double-hit” hypothesis is important to test, as it makes significant claims that would potentially alter both policy and treatment for future pandemics. The hypothesis would, for example, provide an additional justification for government-mandated stay-at-home orders, which are associated with short-term reductions in air pollution by lowering traffic and industrial activity. Previous workers have suggested that decreases in air pollution following the quarantine periods may have also decreased the total number of COVID-19 deaths during those periods [6].

Several ecological studies have presented evidence for an association between air pollution and increased COVID-19 mortality. In Europe as of March 19, 2020, 83% of all COVID-19 fatalities (n = 3,701) occurred in regions where the maximum NO_2_ concentration was above 100 µmol/m² and only 1.5% of all COVID-19 fatalities occurred where NO_2_ concentration was below 50 µmol/m² [2]. An ecological regression analysis of all counties in the United States used a negative binomial mixed model with daily county-level COVID-19 death counts through 18 June 2020 and surface level PM_2.5_ data from 2000 through 2016 to estimate the effect of PM_2.5_ on COVID-19 mortality. The model estimated that an increase of 1 µg/m³ in average PM_2.5_ is associated with a statistically significant 11% (95% CI, 6% to 17%) increase in a county’s COVID-19 mortality rate [3].

Subsequent evidence has remained mixed. A systematic review of 116 studies found positive and statistically significant associations in 48.1% of the reported mortality estimates; all of the studies were observational and most had a high risk of confounding and outcome-measurement bias [7]. A nationwide cohort study of 3.7 million Danish adults reported positive associations between long-term PM_2.5_ and NO_2_ exposure and COVID-19 hospitalization and death [8]. These later studies provide stronger evidence that an association may exist, but they do not eliminate the problem of spatial confounding or establish that the double-hit mechanism explains COVID-19 mortality.

The air pollution hypothesis makes the prediction that particulate and NO_2_ pollution both have positive correlations with COVID-19 mortality. This makes it reasonable to test the hypothesis by comparing these pollution measures with deaths. But there are difficulties with doing this directly.

First, air pollution in cities can be tightly linked to location and varies both spatially and temporally. This creates a problem when pollution and mortality data are not measured at comparable scales. We reduce this problem by linking the times and locations in individual death records with modeled and observed PM_2.5_ and NO_2_ data, while recognizing that the assigned values remain spatial exposure proxies.

Second, air pollution might be acting as a proxy for an unobserved variable, such as demography, poverty, mobility, population density, or housing type [9]. These associated variables could well be the real cause of any observed correlation, which would be misleading as air quality would then have a correlative but not causative relationship with COVID-19 mortality [10].

There is a way around some of this confounding, as the COVID-19 mortality/air-quality hypothesis makes another, implicit prediction: air quality should be more important for COVID-19 mortality than for non-COVID mortality. Comparing COVID and non-COVID natural deaths from the same data source and study period places both groups under the same broad spatial and pandemic conditions. Although air quality has well-established effects on overall mortality [11, 12, 13], the proposed double-hit mechanism predicts an additional association with COVID-19 mortality. If that association is not stronger for COVID deaths, then the observed pollution pattern would not provide evidence specific to the proposed double-hit mechanism and could instead reflect other factors associated with air pollution and all-cause mortality.

The existing research and ecological studies that associate air pollution levels with COVID-19 outcomes have been criticized as lacking rigor [14, 10] because they rely on COVID-19 summary data that is not age-adjusted and use air pollution data that is not spatially or temporally aligned. These studies are not definitive because they have short durations (sometimes only covering one month of sample data – [15, 16]), small sample sizes [2], and may not account for important confounders [17, 18, 19, 20].

To help resolve these issues, we compare modeled and observed measures of PM_2.5_ and nitrogen dioxide among COVID and non-COVID natural deaths in Cook County, Illinois. By linking Cook County Medical Examiner records with pollution estimates, we can test whether air pollution distinguishes COVID from non-COVID deaths after accounting for age, sex, recorded race, and Latino ethnicity. The detailed times and locations in the death records also allow us to test whether short-term NO_2_ associations reflect unusually high exposure before death or are better explained by where deaths occurred.

## Materials and methods

We used two complementary approaches to test whether pollution exposure patterns differed for COVID versus non-COVID deaths: a logistic regression comparing COVID and non-COVID deaths, and a Monte Carlo simulation comparing observed pollution exposure at death locations with an age-adjusted, population-weighted expectation.

### Data sources and processing

A retrospective study was conducted using natural-cause death records from the Cook County Medical Examiner’s Office. The primary analysis compared records classified as COVID and non-COVID deaths occurring during the same period, from March 2020 through August 2021. This contemporaneous comparison was used so that both groups experienced the same broad secular and pandemic-period conditions. To address the possibility that some contemporaneous deaths classified as non-COVID were undiagnosed COVID deaths, we repeated the same analysis using non-COVID natural deaths from 2015–2019 as a robustness check. Logistic regression was used to determine whether demographic characteristics and modeled long-term PM_2.5_ exposure distinguished COVID from non-COVID death records within the Medical Examiner’s sample [21, 22].

The Cook County Medical Examiner’s records contain incident-location information, including latitude and longitude coordinates and incident address fields. The analysis used the coordinates present in the processed records. These coordinates represent the location associated with the death record and do not necessarily represent the person’s primary residence. We therefore interpret pollution assigned at these coordinates as a spatial exposure proxy at the incident or death location rather than as a complete residential exposure history.

The Medical Examiner’s file contains demographic information for each death, including exact age at death, gender, race, and ethnicity. Ages were binned into ranges for comparison with Census tables and vital statistics reports, and the gender, race, and ethnicity fields were one-hot encoded to True/False values for ingestion into statistical tests.

Causes of death are listed in freeform text fields and are subject to misspellings and inconsistent transcription. Regular expressions were used to identify mentions of common disease categories across the primary and secondary cause fields. The COVID classification was derived from these same fields. Consequently, the disease indicators describe conditions mentioned in the death record rather than independently ascertained pre-existing conditions. They were excluded from the primary adjusted model to avoid using variables derived from the outcome source as baseline covariates. Analyses using these disease mentions and incident-setting variables are treated separately as descriptive classification of Medical Examiner records and are not interpreted as estimates of pre-existing disease risk.

### PM2.5 data WUSTL GEOS Chem model

The Atmospheric Composition Analysis Group at Washington University in St. Louis provides gridded estimates of surface-level fine particulate matter by combining satellite remote sensing with the GEOS-Chem chemical transport model and calibrating the estimates to ground-based observations [23, 24]. We used the V5.GL.02 monthly North American PM_2.5_ product from January 2000 through December 2020. The estimates are gridded at 0.01° × 0.01°, which in Cook County equates to cells roughly 1.1 km north-to-south by 0.8 km east-to-west. Each death was assigned the value for the grid cell containing its recorded coordinates, and long-term exposure for each look-back interval was calculated as the mean of the available monthly estimates [25].

To compare average PM_2.5_ exposure between persons who died from COVID-19 and the general population, an areal interpolation of the PM_2.5_ surface estimates to Census block group was conducted, because a simple spatial mean gives equal weight to unpopulated or less dense areas and so underestimates population exposure. The geographic centroid of each of the 3,993 Cook County Census block groups was assigned to the appropriate grid cell; this is efficient because block groups are small relative to grid cells and the PM_2.5_ gradient between cells is not steep within the study area [26] (Supplementary Figure S1).

### NO2 data EPA CMAQ model and AQS monitors

The Community Multiscale Air Quality (CMAQ) modeling system is an open-source EPA project that provides hourly estimates of pollutant concentrations on the WRF 12 km simulation grid. In response to pandemic changes in activity and emissions, EPA’s Office of Research and Development produced a specialized CMAQ model for the continental United States that adjusts the base model using INRIX vehicle-miles-traveled data [27]. The retained hourly CMAQ input covered January 1 through July 31, 2020 (local conversion begins December 31, 2019), while deaths were drawn from the March–July first-wave period. The grid was sliced to Cook County and the appropriate cell assigned to each death. To keep the centered ±14-day randomization window complete, deaths later than the maximum CMAQ timestamp minus 14 days were excluded. The January start provided complete coverage for the longest 168-hour look-back and the lower randomization boundary [28].

The Air Quality System (AQS) is EPA’s national network of ground monitoring stations. In Cook County, the only urban-scale NO_2_ monitor (site #4201) was offline for most of 2020. Two neighborhood-scale monitors (sites #76 and #4002), each with a 4,000 m measurement scale, are the source of hourly observational NO_2_ data used here; observational data were restricted to residents within 4,000 m of a station, consistent with the calibrated monitoring scale. The remaining sites are near “trouble spots” (O’Hare, major highways) and are not independently and identically distributed, so CMAQ was used for county-wide NO_2_ estimates rather than interpolating those sites [29].

The AQS NO_2_ series showed strong diurnal, weekly, and seasonal structure, which informed the temporal randomization analysis.

### Logistic model

The logistic model was used to test whether modeled long-term PM_2.5_ exposure distinguished records classified as COVID deaths from contemporaneous non-COVID natural deaths after accounting for demographic differences. The response variable indicated whether COVID was recorded in the cause-of-death fields. Predictors were age band, sex, recorded race, Latino ethnicity, and the modeled 2020 five-year mean PM_2.5_ concentration assigned to the incident or death location. Ages were divided into eight bands (<20, 20–29, 30–39, 40–49, 50–59, 60–69, 70–79, and ≥80 years); reference categories were 50–59 years, male sex, White race, and not Latino. The PM_2.5_ odds ratio was estimated per 1 µg/m³. For context, we also report the same coefficient per pooled within-group SD of 0.303 µg/m³; this is an algebraic rescaling, not a refitted model.

Models were fitted to complete cases using observed records, an intercept, and no resampling or feature selection. For records with non-missing PM_2.5_ exposure data, only 17 were excluded because of incomplete demographic covariates (8 COVID and 9 non-COVID; ∼0.1% of records). The contemporaneous primary model included 15,010 deaths (10,887 COVID and 4,123 non-COVID). The sensitivity model used the same predictors and COVID cases but substituted non-COVID natural deaths from 2015–2019, yielding 22,935 complete records (10,887 COVID and 12,048 non-COVID). Results are reported as odds ratios with 95% confidence intervals and two-sided p-values; model fit is summarized using McFadden’s pseudo-R². Mean PM_2.5_ exposure was also compared directly between COVID and non-COVID deaths using a two-sided Welch t-test. Analysis-specific cohorts, date windows, exposure coverage, and denominators are summarized in Supplementary Table S1. Denominators differ because exposure availability and complete-case requirements differ among analyses.

### Monte Carlo simulation of exposure

We used a series of Monte Carlo analyses to compare observed pollution exposure in each death group with the expected exposure of its age-adjusted countywide population. Since Monte Carlo simulations approximate the full range and likelihood of possible outcomes, the results are superior to single-point estimates for risk assessments [30]. This approach requires us to spatially and temporally weight the population to match the demographic characteristics of the corresponding death group.

We used the Cook County Medical Examiner’s data to construct a probabilistic age distribution for each death group. We then built the spatial probability distribution using areal interpolation of Census block groups to the grid cells in our air quality data, assigning a pro rata population proportion by age range to each grid cell.

A series of Monte Carlo analyses was conducted for each death group to compare actual NO_2_ exposure at various time intervals in the week prior to death versus hypothetical exposure using a randomly selected hour of death occurring in a 28-day period centered at the actual time of death. The NO_2_ data contain hourly observations for each cell, allowing us to estimate mean exposure in the 24, 48, 72, 96, 120, 144, and 168 hours prior to death. The inclusion of temporal randomness allows us to account for diurnal variability in NO_2_.

Comparing expected and observed PM_2.5_ is somewhat simpler, as PM_2.5_ does not have the periodic variability of NO_2_, and so only requires spatial weighting. The PM_2.5_ data contain monthly observations for each cell, allowing us to use the Monte Carlo analyses to estimate the mean exposure of each death group in the 12, 24, 36, 48, 60, 72, 84, 120, 180, and 240 months prior to death, which is then compared with the corresponding weighted population for the same period.

For deaths occurring after December 2020, WUSTL look-back means used the available monthly estimates through December 2020; unavailable later months were excluded from both the observed and simulated means.

We also calculated hourly average NO_2_ values for 2020 and 2021 from EPA site #4002. Annual AQS PM_2.5_ values were calculated from the available Cook County-area monitoring data and population-weighted by the monitoring areas used in the spatial workflow. The retained annual AQS series contains no values for 2011– 2013. The corresponding WUSTL series gives population-weighted annual means across Cook County (Figure 1).

**Figure 1.**
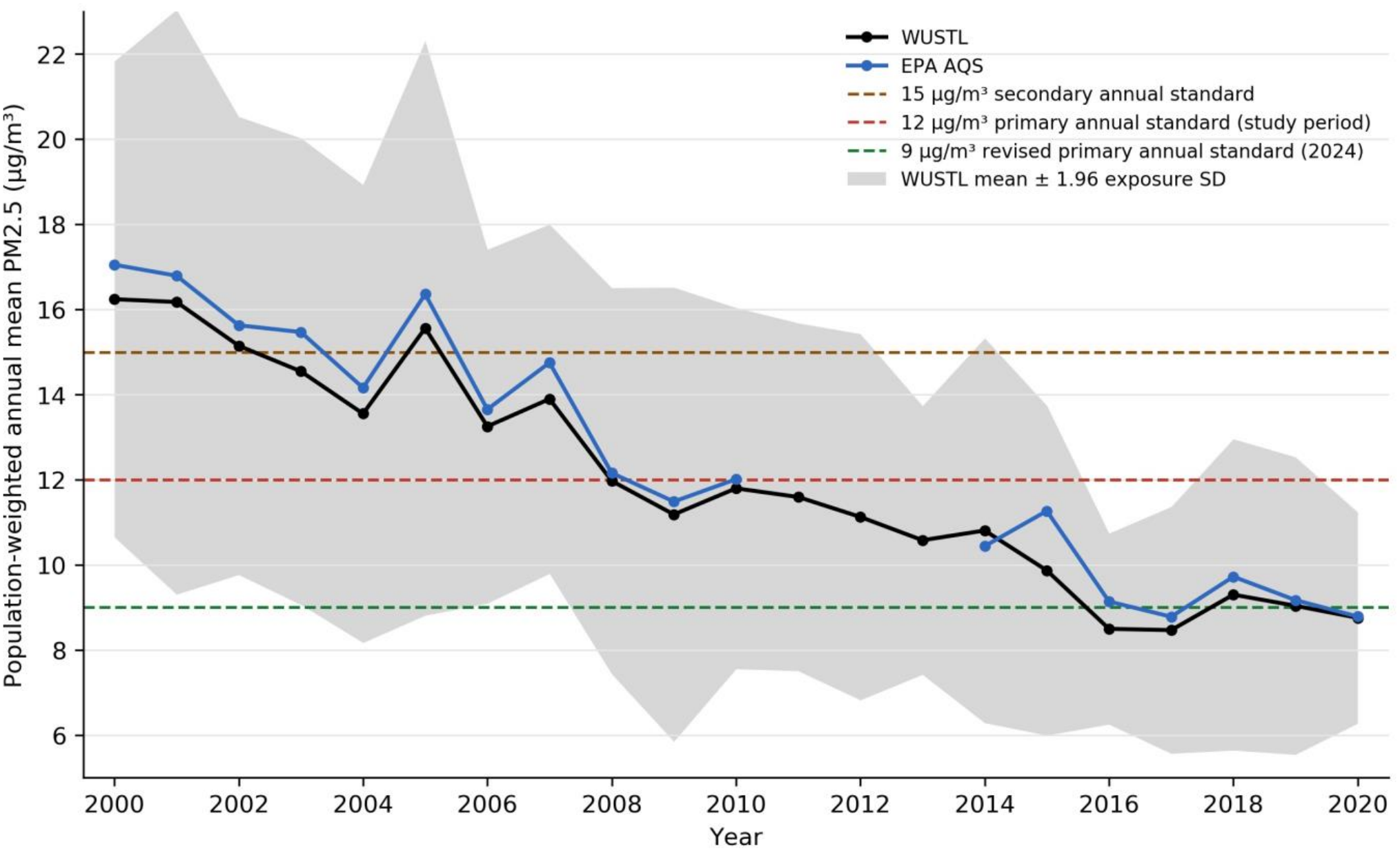
Population-weighted mean PM_2.5_ exposure in Cook County, 2000–2020. Black points and lines show WUSTL estimates; blue points and lines show the population-weighted AQS aggregation across available monitoring areas. The gray ribbon shows the WUSTL annual mean ± 1.96 population-weighted SD across the monthly grid-cell exposure distribution (an approximate 95% exposure band, not a confidence interval for the mean). The retained AQS annual series has no values for 2011–2013. Horizontal lines show the 15 µg/m³ secondary annual standard, the 12 µg/m³ primary annual standard applicable during the study period, and the revised 9 µg/m³ primary annual standard adopted in 2024 [31].

We used a simulated sample size equal to the number of deaths in the relevant group and conducted each Monte Carlo analysis with 10,000 trials to provide a practical balance between computational efficiency and stable estimation. Locations were drawn from the age-adjusted, population-weighted spatial distribution; the NO_2_ analyses also randomized time by drawing an hour from a uniform 28-day window centered on the actual time of death, whereas the PM_2.5_ analyses required only spatial randomization. Since the hypothesis predicts higher exposure, we calculated the empirical right-tail p-value as the proportion of simulated means at least as large as the observed mean, and calculated the Z-score as the difference between the observed and expected means divided by the standard deviation of the simulated means.

For the NO_2_ robustness analysis, observed exposure at the recorded death locations and times was compared with two simulated reference distributions. First, reference locations were randomized using population weights while each death’s timing was retained, testing the spatial exposure contrast at the observed death times. Second, reference times were randomized within the centered 28-day window while each death’s location was retained, testing whether exposure immediately before death was unusually high relative to nearby times at the same locations.

ChatGPT was used to assist with manuscript editing, formatting, and checks of internal consistency.

## Results

### Logistic model

Using natural-cause deaths occurring during the COVID-19 study period as the comparison group, mean five-year PM_2.5_ exposure was slightly lower among COVID-19 deaths than among non-COVID deaths (difference = −0.049 µg/m³, 95% CI −0.059 to −0.039; two-sided Welch p = 1.12 × 10⁻²¹). In the primary adjusted model containing only variables measured independently of the cause-of-death text—age, sex, recorded race, Latino ethnicity, and modeled PM_2.5_ exposure—higher PM_2.5_ was associated with lower odds that a Medical Examiner record was classified as a COVID rather than a non-COVID death (OR = 0.63 per 1 µg/m³, 95% CI 0.55–0.72; p < 0.001). The same coefficient corresponds to OR = 0.87 (95% CI 0.83–0.91) per pooled within-group SD of 0.303 µg/m³. As a robustness check addressing possible undiagnosed COVID deaths in the contemporaneous comparison group, we repeated the same analysis using natural-cause deaths from 2015–2019. The association remained negative (OR = 0.54, 95% CI 0.48–0.60; p < 0.001) (Table 1).

**Table 1.** Demographic characteristics and modeled five-year PM_2.5_ exposure associated with classification as a COVID rather than non-COVID natural death among Cook County Medical Examiner records, March 2020–August 2021.

| Explanatory variable | Adjusted OR (95% CI) | p-value | Non-COVID | COVID |
|---|---|---|---|---|
| **Age range** |  |  |  |  |
| <20 | 0.32 (0.17, 0.58) | <0.001 | 38 (0.9%) | 17 (0.2%) |
| 20–29 | 0.50 (0.35, 0.73) | <0.001 | 84 (2.0%) | 57 (0.5%) |
| 30–39 | 0.63 (0.51, 0.78) | <0.001 | 246 (6.0%) | 200 (1.8%) |
| 40–49 | 0.77 (0.65, 0.91) | 0.002 | 472 (11.4%) | 483 (4.4%) |
| 50–59 | Reference | — | 859 (20.8%) | 1,049 (9.6%) |
| 60–69 | 1.54 (1.36, 1.73) | <0.001 | 1,280 (31.0%) | 2,194 (20.2%) |
| 70–79 | 3.61 (3.18, 4.11) | <0.001 | 691 (16.8%) | 2,745 (25.2%) |
| ≥80 | 8.18 (7.11, 9.40) | <0.001 | 453 (11.0%) | 4,142 (38.0%) |
| **Sex** |  |  |  |  |
| Male | Reference | — | 2,886 (70.0%) | 6,214 (57.1%) |
| Female | 1.50 (1.38, 1.63) | <0.001 | 1,237 (30.0%) | 4,673 (42.9%) |
| **Recorded race** |  |  |  |  |
| White | Reference | — | 2,180 (52.9%) | 6,805 (62.5%) |
| Black | 0.99 (0.91, 1.08) | 0.827 | 1,783 (43.2%) | 3,190 (29.3%) |
| Asian | 1.74 (1.37, 2.22) | <0.001 | 98 (2.4%) | 447 (4.1%) |
| Other | 2.87 (1.96, 4.20) | <0.001 | 34 (0.8%) | 275 (2.5%) |
| AI/AN | 0.58 (0.16, 2.09) | 0.410 | 5 (0.1%) | 6 (0.1%) |
| Unknown | 2.36 (1.48, 3.77) | <0.001 | 23 (0.6%) | 164 (1.5%) |
| **Latino ethnicity** |  |  |  |  |
| Not Latino | Reference | — | 3,693 (89.6%) | 8,539 (78.4%) |
| Latino | 4.03 (3.54, 4.58) | <0.001 | 430 (10.4%) | 2,348 (21.6%) |
| **Modeled PM2.5 exposure** |  |  |  |  |
| 2020 five-year mean, per 1 µg/m³ | 0.63 (0.55, 0.72) | <0.001 | 8.853 (0.268) | 8.804 (0.315) |

Notes: Values in the Non-COVID and COVID columns are observed n (column %) for categorical variables and mean (SD) for PM<sub>2.5</sub>. The model includes 15,010 observed natural deaths: 4,123 non-COVID and 10,887 COVID deaths. Odds ratios estimate the odds that a Medical Examiner record was classified as COVID rather than non-COVID within this sample; they do not estimate population-level mortality risk. Reference categories are age 50–59 years, male sex, White race, and not Latino. AI/AN denotes American Indian or Alaska Native; the AI/AN estimate is based on a small sample size and should be interpreted cautiously. The PM<sub>2.5</sub> odds ratio is reported per 1 µg/m³; the same coefficient corresponds to OR = 0.87 (95% CI 0.83–0.91) per pooled within-group SD of 0.303 µg/m³. The fixed five-year PM<sub>2.5</sub> exposure ends in 2020. Exposure is a modeled spatial proxy assigned at the incident/death location, not verified residential history. The model includes an intercept and uses observed records only. McFadden pseudo-R² = 0.148.

### Monte Carlo simulation of exposure

The modeled WUSTL PM_2.5_ and CMAQ NO_2_ analyses showed exposures above the respective population-weighted expectations for COVID-19 deaths. However, similar analyses for non-COVID deaths showed equal or larger departures from expectation, indicating that the pattern was not specific to COVID-19. Both COVID and non-COVID deaths were associated with locations above their population-weighted WUSTL expectations at all examined time periods (Figure 2, Table 2). The absolute observed-minus-expected differences were small (approximately 0.02–0.14 µg/m³), but the large group sizes produced tight simulated distributions of the group means and WUSTL z-scores above 5. The smaller AQS data set showed a less consistent pattern.

**Figure 2.**
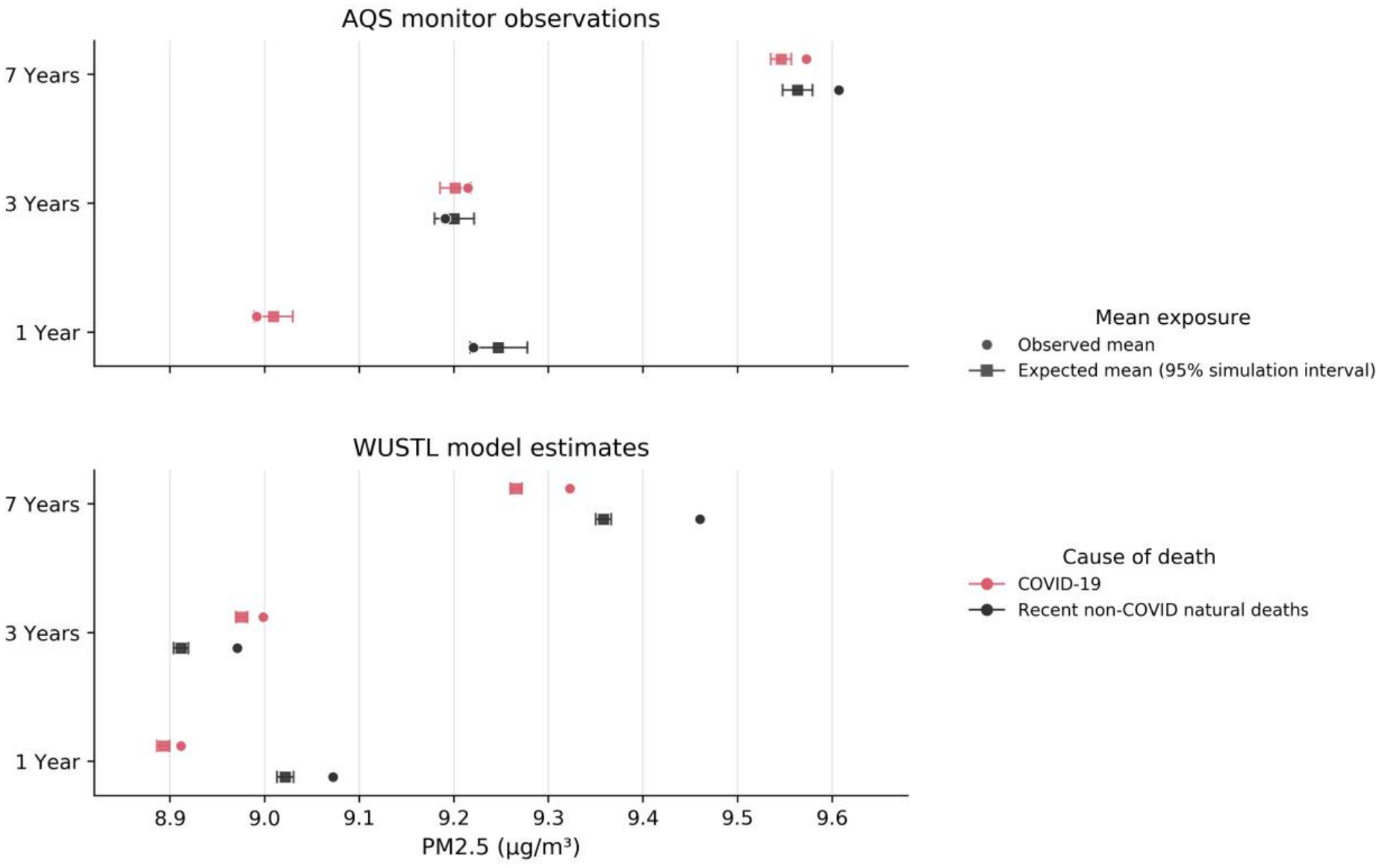
Observed and population-weighted expected mean PM_2.5_ exposure for COVID-19 deaths and recent non-COVID natural deaths over nominal death-relative 1-, 3-, and 7-year look-back periods. The top panel uses AQS monitor observations and the bottom panel uses WUSTL model estimates. Circles show observed means; squares and horizontal bars show expected means and central 95% intervals from 10,000 Monte Carlo simulations. For deaths after December 2020, WUSTL look-back means use the available monthly estimates through December 2020. Sample sizes were 3,374 COVID and 1,831 recent non-COVID deaths for AQS, and 11,850 and 7,548, respectively, for WUSTL; the recent non-COVID group spans December 31, 2018 through December 31, 2021.

**Table 2.** Observed versus population-weighted expected PM_2.5_ exposure prior to death, with central 95% simulation intervals and empirical right-tail p-values from 10,000 Monte Carlo trials.

| Group | Exposure (months) | n | Observed mean ± SD | Expected mean (95% simulation interval) | Empirical right-tail p |
|---|---|---|---|---|---|
| COVID-19 | 12 | 11,850 | 8.91 ± 0.31 | 8.893 (8.886, 8.899) | <0.0001 |
| COVID-19 | 24 | 11,850 | 9.07 ± 0.33 | 9.048 (9.041, 9.054) | <0.0001 |
| COVID-19 | 36 | 11,850 | 9.00 ± 0.29 | 8.976 (8.970, 8.982) | <0.0001 |
| COVID-19 | 48 | 11,850 | 8.89 ± 0.30 | 8.852 (8.846, 8.858) | <0.0001 |
| COVID-19 | 60 | 11,850 | 8.87 ± 0.32 | 8.819 (8.812, 8.825) | <0.0001 |
| COVID-19 | 72 | 11,850 | 9.06 ± 0.35 | 9.003 (8.997, 9.010) | <0.0001 |
| COVID-19 | 84 | 11,850 | 9.32 ± 0.35 | 9.266 (9.260, 9.272) | <0.0001 |
| COVID-19 | 120 | 11,850 | 9.89 ± 0.36 | 9.832 (9.825, 9.839) | <0.0001 |
| COVID-19 | 180 | 11,850 | 10.83 ± 0.41 | 10.773 (10.766, 10.780) | <0.0001 |
| COVID-19 | 240 | 11,850 | 11.87 ± 0.46 | 11.799 (11.791, 11.808) | <0.0001 |
| Recent non-COVID natural deaths | 12 | 7,548 | 9.07 ± 0.37 | 9.022 (9.013, 9.030) | <0.0001 |
| Recent non-COVID natural deaths | 24 | 7,548 | 9.04 ± 0.30 | 8.985 (8.977, 8.993) | <0.0001 |
| Recent non-COVID natural deaths | 36 | 7,548 | 8.97 ± 0.29 | 8.912 (8.904, 8.919) | <0.0001 |
| Recent non-COVID natural deaths | 48 | 7,548 | 8.92 ± 0.29 | 8.846 (8.838, 8.854) | <0.0001 |
| Recent non-COVID natural deaths | 60 | 7,548 | 9.01 ± 0.34 | 8.923 (8.915, 8.931) | <0.0001 |
| Recent non-COVID natural deaths | 72 | 7,548 | 9.22 ± 0.39 | 9.124 (9.116, 9.132) | <0.0001 |
| Recent non-COVID natural deaths | 84 | 7,548 | 9.46 ± 0.38 | 9.358 (9.350, 9.366) | <0.0001 |
| Recent non-COVID natural deaths | 120 | 7,548 | 10.03 ± 0.38 | 9.920 (9.912, 9.929) | <0.0001 |
| Recent non-COVID natural deaths | 180 | 7,548 | 11.03 ± 0.45 | 10.911 (10.901, 10.920) | <0.0001 |
| Recent non-COVID natural deaths | 240 | 7,548 | 11.90 ± 0.41 | 11.759 (11.746, 11.772) | <0.0001 |

Notes: Values are PM<sub>2.5</sub> concentrations in µg/m³. The exposure labels are nominal death-relative monthly look-back windows. The empirical p-value is the proportion of 10,000 simulated means at least as large as the observed mean. A displayed value of <0.0001 means that no simulated mean equaled or exceeded the observed mean. These separate null comparisons do not constitute a direct COVID-versus-non-COVID test.
For deaths occurring after December 2020, the nominal look-back windows used available WUSTL monthly estimates through December 2020; unavailable later months were excluded from both observed and simulated means.
The recent non-COVID comparison is a negative-comparison group comprising natural deaths dated December 31, 2018 through December 31, 2021 (n = 7,548); it is distinct from the primary contemporaneous logistic comparison group. Because this window overlaps the pandemic period, some records classified as non-COVID could include undiagnosed COVID deaths.

The long-term decline in expected PM_2.5_ concentrations (Table 2) is the result of a secular decline in PM_2.5_ across the study domain (Figure 1). The population-weighted WUSTL annual mean fell from 16.2 µg/m³ in 2000 to 8.8 µg/m³ in 2020, and the 2020 mean was below the lower bound of the approximate 95% exposure band for 2000. The population-weighted AQS series closely tracked WUSTL where available; the retained AQS annual series has no values for 2011–2013. The 12 µg/m³ annual primary standard shown for the study period was revised to 9 µg/m³ in 2024 [31].

The relationship between deaths and short-term NO_2_ exposure shows less of a pattern than that shown in the PM_2.5_ case (Figure 3). For COVID deaths, the AQS point estimates did not exceed the upper bound of the corresponding central 95% simulation interval at any time frame. Some AQS comparisons for non-COVID natural deaths did exceed the interval, including the 24-hour mean and the mean and maximum values for the 48-through 168-hour periods. The county-wide CMAQ results showed modest but statistically detectable excess exposure for COVID deaths across the 24-through 168-hour periods (mean differences of 0.13–0.19 ppb; empirical p = 0.043 to <0.001). However, excesses were also present, and were consistently larger, for contemporaneous non-COVID natural deaths (0.82–0.99 ppb; all empirical p < 0.0001) (Table 3).

**Figure 3.**
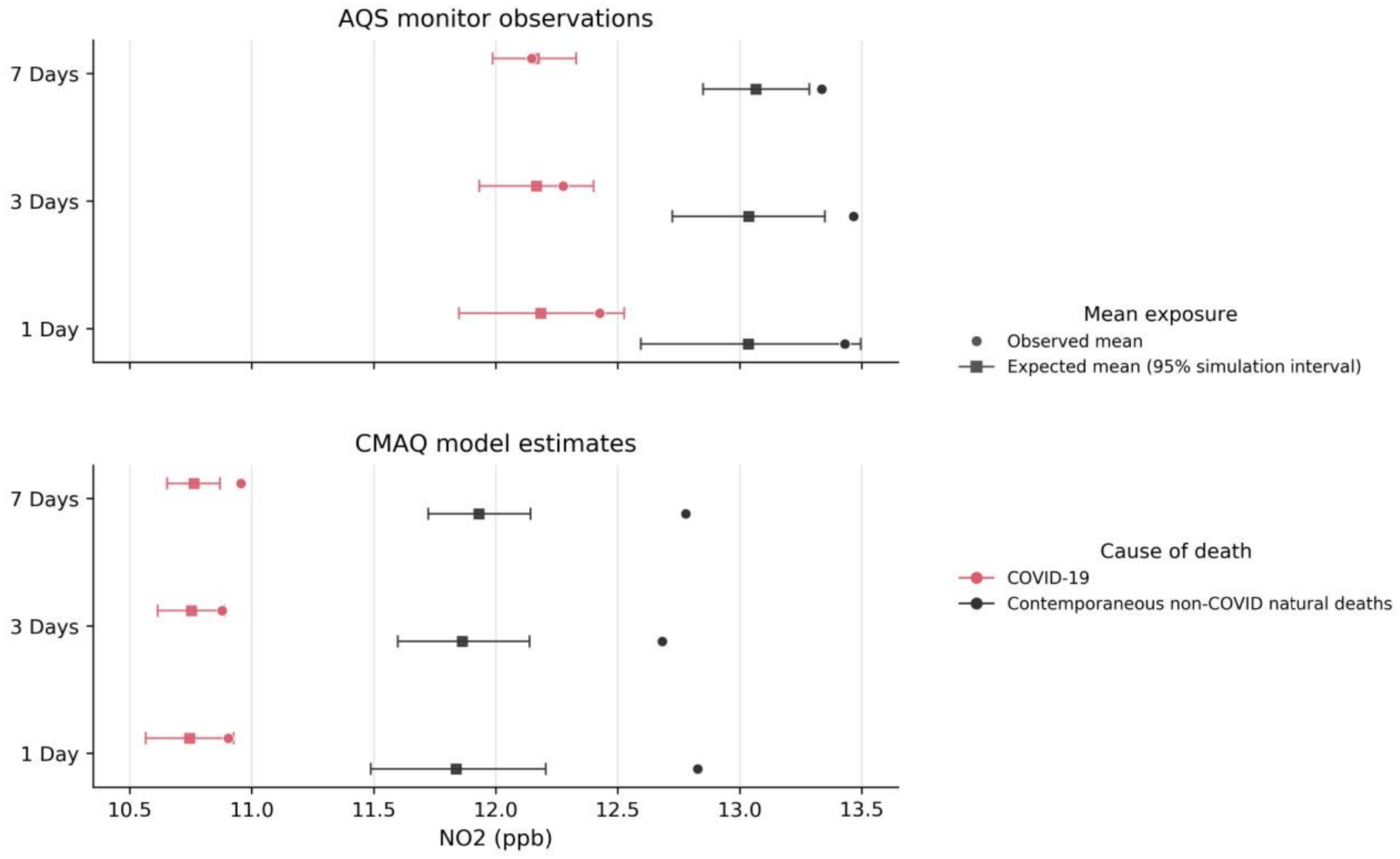
Observed and population-weighted expected mean NO_2_ exposure for COVID and non-COVID deaths over 1-, 3-, and 7-day look-back periods. The top panel uses available AQS monitor observations from 2015–2021, with eligible deaths restricted by monitor coverage and complete centered time windows; the bottom panel uses CMAQ model estimates for the 2020 first-wave period. Circles show observed means; squares and horizontal bars show expected means and central 95% intervals from 10,000 Monte Carlo simulations. Sample sizes were 1,206 COVID and 659 non-COVID deaths for AQS, and 4,522 COVID and 1,377 contemporaneous non-COVID natural deaths for CMAQ.

**Table 3.** Observed versus population-weighted expected NO_2_ exposure before death, by death group and exposure window.

| **Exposure (hours)** | **Mean NO2, observed** | **Mean NO2, expected (95% simulation interval)** | **Empirical p** | **1-hour maximum NO2, observed** | **1-hour maximum NO2, expected (95% simulation interval)** | **Empirical p** |
|---|---|---|---|---|---|---|
| **COVID-19 (n=4,522)** |  |  |  |  |  |  |
| 24 | 10.9 ± 5.7 | 10.7 (10.6, 10.9) | 0.0429 | 24.0 ± 11.5 | 23.5 (23.1, 23.8) | 0.0037 |
| 48 | 10.9 ± 4.8 | 10.7 (10.6, 10.9) | 0.0148 | 29.7 ± 10.3 | 29.1 (28.8, 29.4) | 0.0001 |
| 72 | 10.9 ± 4.2 | 10.8 (10.6, 10.9) | 0.0341 | 32.9 ± 9.2 | 32.4 (32.1, 32.7) | 0.0002 |
| 96 | 10.9 ± 3.8 | 10.8 (10.6, 10.9) | 0.0229 | 34.9 ± 8.6 | 34.4 (34.2, 34.7) | 0.0004 |
| 120 | 10.9 ± 3.5 | 10.8 (10.6, 10.9) | 0.0113 | 36.3 ± 8.2 | 35.9 (35.7, 36.2) | 0.0010 |
| 144 | 10.9 ± 3.4 | 10.8 (10.6, 10.9) | 0.0019 | 37.5 ± 7.9 | 37.1 (36.8, 37.4) | 0.0021 |
| 168 | 11.0 ± 3.2 | 10.8 (10.7, 10.9) | 0.0001 | 38.5 ± 7.7 | 38.1 (37.8, 38.3) | 0.0011 |
| **Contemporaneous non-COVID natural deaths (n=1,377)** |  |  |  |  |  |  |
| 24 | 12.8 ± 7.3 | 11.8 (11.5, 12.2) | <0.0001 | 26.6 ± 13.0 | 24.8 (24.2, 25.5) | <0.0001 |
| 48 | 12.7 ± 6.1 | 11.8 (11.5, 12.2) | <0.0001 | 32.0 ± 11.4 | 30.4 (29.8, 31.0) | <0.0001 |
| 72 | 12.7 ± 5.5 | 11.9 (11.6, 12.1) | <0.0001 | 35.2 ± 10.4 | 33.7 (33.1, 34.2) | <0.0001 |
| 96 | 12.8 ± 5.1 | 11.9 (11.6, 12.1) | <0.0001 | 37.4 ± 10.0 | 35.8 (35.3, 36.4) | <0.0001 |
| 120 | 12.8 ± 4.9 | 11.9 (11.7, 12.1) | <0.0001 | 38.8 ± 9.5 | 37.4 (36.9, 37.9) | <0.0001 |
| 144 | 12.8 ± 4.7 | 11.9 (11.7, 12.1) | <0.0001 | 40.1 ± 9.1 | 38.6 (38.2, 39.1) | <0.0001 |
| 168 | 12.8 ± 4.5 | 11.9 (11.7, 12.1) | <0.0001 | 41.0 ± 8.9 | 39.6 (39.2, 40.1) | <0.0001 |

Notes: NO<sub>2</sub> values are in ppb and are shown as mean ± SD. Expected values and central 95% simulation intervals are from 10,000 population-weighted Monte Carlo trials. Empirical right-tail p-values are the proportions of simulated means at least as large as the observed mean; <0.0001 means that no simulated mean equaled or exceeded the observed mean. The group-specific null comparisons are not a direct statistical comparison between COVID-19 and non-COVID deaths.

### Temporal variability

NO_2_ is periodic in the data at the daily, weekly and seasonal scales, as is observed in the AQS tower data. The average daily high occurs at 6am and is 19.9 ± 11.1 ppb; the average daily low occurs at 2pm and is 8.5 ± 5.1 ppb. Weekdays have higher values (14.6 ± 9.8 ppb) than weekends (12.0 ± 9.4 ppb). There is also a strong seasonal component, with winter values averaging 18.1 ± 11.0 ppb and summer values averaging 11.3 ± 8.2 ppb. These patterns describe general temporal variability in NO_2_; the temporal randomization analysis separately tested whether exposure immediately before death was unusually high relative to nearby times at the same locations.

The AQS observations show the same broad temporal pattern as WUSTL, with population-weighted annual means declining from 17.1 µg/m³ in 2000 to 8.8 µg/m³ in 2020 (Figure 1).

Two robustness checks indicate that the pollution results are not evidence of an effect specific to COVID deaths. First, the negative association between five-year PM_2.5_ exposure and classification as a COVID rather than non-COVID natural death was retained when the comparison group was restricted to natural-cause deaths from 2015–2019, eliminating possible contamination by undiagnosed COVID deaths in the contemporaneous comparison group (OR = 0.54, 95% CI 0.48–0.60; p < 0.001). Second, observed CMAQ NO_2_ exposure exceeded the reference exposure obtained by randomizing locations while retaining each death’s timing (mean-exposure z-scores 2.70–5.20 across the 24-through 168-hour periods). When reference times were randomized while each death’s location was retained, observed mean exposure showed no positive excess (z = −1.80 to −0.44). These comparisons support a spatial exposure contrast rather than unusually high NO_2_ exposure immediately before death.

## Discussion

### Logistic model

The logistic model found that age was the clearest demographic factor distinguishing COVID from non-COVID natural deaths. The odds that a record was classified as COVID increased steadily with age, and were also higher for women and Latino decedents (Table 1). This is consistent with previous work showing that age and other demographic characteristics were important predictors of COVID-19 mortality [32]. It should be noted that our analysis differs from the earlier study in that it compares COVID and non-COVID natural deaths: it does not estimate the population-level risk of dying from COVID-19.

In contrast, our results did not find evidence that long-term PM_2.5_ exposure distinguished COVID from non-COVID natural deaths in the direction predicted by the double-hit hypothesis. The adjusted relationship was negative in both the contemporaneous analysis and the pre-pandemic robustness check. This does not imply that PM_2.5_ exposure is protective. Rather, within this sample, long-term PM_2.5_ was not more strongly associated with COVID deaths than with other natural deaths. This argues against the “double-hit” hypothesis, which privileges the role of PM_2.5_ in COVID-19 mortality.

### Monte Carlo simulation of exposure

If the double-hit hypothesis is true, we would expect higher than expected long-term PM_2.5_ exposure and higher than expected short-term exposures to NO_2_. Modeled PM_2.5_ exposure values are higher than expected for COVID-19 deaths, with very high Z-scores regardless of which long-term time frame is considered most important. Considered alone, this is consistent with the double-hit hypothesis. But the same pattern occurs among non-COVID deaths, and the direct comparison found slightly lower five-year PM_2.5_ exposure among COVID deaths. Thus, particulate exposure does not have the stronger relationship with COVID deaths that the hypothesis predicts.

The NO_2_ exposure analysis is also not consistent with the double-hit hypothesis. The AQS observations did not show excess exposure among COVID deaths. The CMAQ model found a small positive excess for COVID deaths, but the excess was consistently larger for non-COVID deaths. These simulations compare each group with its own population-weighted expectation and are not a direct statistical test between the groups. We therefore treat the difference in excesses as descriptive rather than as a formal between-group test; even so, the pattern is not specific to COVID deaths.

The CMAQ robustness analysis helps distinguish spatial exposure contrasts from acute temporal elevations. At the observed death times, NO_2_ exposure at death locations exceeded the population-weighted expectation from randomized reference locations. At the observed death locations, however, exposure immediately before death did not exceed the expectation from randomized nearby reference times. The findings therefore support a spatial interpretation of the association and provide no evidence of unusually elevated NO_2_ exposure immediately before COVID deaths under this comparison. They do not establish whether the spatial association reflects pollution exposure, other characteristics of the death locations, or both.

The mortality associations with PM_2.5_ do not by themselves establish a special COVID-19 mechanism. It appears likely from our data that PM_2.5_ and NO_2_ exposure are spatial proxies for other health impacts. When we look at PM_2.5_ exposure alone we find a strong and consistent connection with mortality, but in the logistic model, after accounting for demographic differences, PM_2.5_ did not distinguish COVID from non-COVID deaths in the predicted direction. This does not imply that air pollution has no health effects. It indicates that the observed pollution associations do not provide evidence for the double-hit hypothesis.

This interpretation is also consistent with the descriptive exposure pattern for deaths not ordinarily attributed to long-term PM_2.5_ exposure: mean five-year PM_2.5_ at locations of accidental, gun-related, and opioid-related deaths was as high as or higher than at COVID death locations (Supplementary Figure S2).

The modeled PM_2.5_ and NO_2_ data (WUSTL and CMAQ) yield consistently stronger exposure/mortality relationships than the AQS observations do (Figures 2, 3). This is not surprising as the modeled area includes the entire domain while the AQS values are restricted to a small fraction of the area, and so capture fewer deaths (Supplementary Figure S1). The AQS analyses were included as observational complements to the spatially complete modeled products. AQS monitor availability resulted in missing data and limited geographic coverage, particularly for NO_2_ (Supplementary Table S2); consequently, the AQS results should be interpreted as sensitivity analyses rather than population-representative exposure estimates. Our experience highlights a challenge for environmental health studies: where the observational data is limited by issues of data quality and geographic coverage, such data lacks sufficient spatial and temporal resolution to find subtle effects.

### Limitations

These data have properties that reduce their precision. Pollution values were assigned to incident or death-record locations and are spatial proxies rather than measurements of personal exposure. Time spent elsewhere, indoor exposure, and residential moves will therefore introduce error, particularly for long-term PM_2.5_ and short-term NO_2_ exposure. This is particularly relevant for NO_2_, as those who died from COVID-19 may have spent a final period of time indoors or in a hospital, where ambient values at the recorded location may not represent actual exposure. The available data for short-term CMAQ NO_2_ analysis was restricted to the first pandemic wave (March–July 2020) and may not generalize to subsequent conditions. The fixed five-year PM_2.5_ measure also varied over a narrow range within Cook County. This constrains interpretation of effect size and generalization to larger exposure contrasts, although the coefficient itself was estimated precisely.

The Medical Examiner’s records are also not a complete sample of deaths in Cook County. The logistic model therefore estimates which characteristics distinguish COVID from non-COVID natural deaths within this sample; it does not estimate the population-level risk of dying from COVID-19. Some contemporaneous non-COVID deaths may have been undiagnosed COVID deaths, although the pre-pandemic robustness check produced the same negative PM_2.5_ relationship. The separate recent non-COVID PM Monte Carlo group (December 31, 2018–December 31, 2021) also overlaps the pandemic period and may include undiagnosed COVID deaths; it is treated as a negative-comparison group rather than the primary exchangeable control group. Finally, the Monte Carlo simulations compare each group with its own population-weighted expectation and are not a direct statistical test between the COVID and non-COVID groups.

We found no evidence that air pollution associations predicted by the “double-hit” hypothesis explained COVID-19 deaths among Cook County Medical Examiner records. Age and demographic characteristics clearly distinguished COVID from non-COVID natural deaths within the Medical Examiner’s records, while long-term PM_2.5_ did not distinguish the groups in the predicted direction and the short-term CMAQ NO_2_ analysis showed a spatial exposure contrast without evidence of unusually high exposure immediately before death. These results do not imply that air pollution has no health effects, but rather reinforce the need for high-quality data in studies that search for acute and long-term air-quality effects. Future studies using verified residential histories, individual-level covariates, population-based or prospective cohorts, or observational air pollution data with greater spatial and temporal coverage could more directly test whether acute and chronic pollution exposures are associated with cause-specific mortality risk.

## Data availability statement

The original death records, population data, and air-quality observations and model outputs are available from the Cook County Medical Examiner, U.S. Census Bureau, U.S. Environmental Protection Agency, and Atmospheric Composition Analysis Group, subject to the access terms of the respective providers. A public repository link for the analysis code and shareable de-identified or aggregated derived data is not provided with this version. The integrated record-level dataset and precise death-location coordinates will not be released.

## Acknowledgments

The authors acknowledge the agencies and research groups that made the Medical Examiner records, Census data, air-quality observations, and modeled exposure products available.

## Author contributions

AF and JT conceived the study. AF collected and curated the data, proposed the analytical tests, performed the original coding and primary analyses, and produced the original figures. JT checked the analyses for errors and internal consistency. AF and JT shared preparation of the original draft and both performed revisions.

## Ethical approval

The University of Illinois Urbana-Champaign Institutional Review Board reviewed this project (IRB26-0878) and determined, on 16 July 2026, that it does not meet the U.S. Department of Health and Human Services definition of human subjects research; IRB oversight was therefore not required. The determination was based on the fact that the study consists solely of the secondary analysis of existing, publicly available data concerning deceased individuals and does not involve obtaining information or identifiable private information about any living individuals. The project also does not meet the U.S. Food and Drug Administration definition of human subjects research.

