## Supplementary information for "COVID specificity of air pollution associations with mortality in Cook County Medical Examiner records"

Preprint supplement prepared 30 October 2026. Not certified by peer review.

### Supplementary Table S1 Analysis cohorts exposure windows and denominators

| Analysis | Purpose and exposure | Date window | Main eligibility rule | COVID n | Non-COVID n |
|---|---|---|---|---|---|
| Direct PM2.5 Welch comparison | Two-sided comparison of the fixed WUSTL five-year mean ending in 2020 | March 1, 2020–August 31, 2021 | Nonmissing PM2.5 | 10,895 | 4,132 |
| Primary logistic model / Table 1 | Classification model using the fixed WUSTL five-year mean ending in 2020 | March 1, 2020–August 31, 2021 | Complete demographic and PM2.5 cases | 10,887 | 4,123 |
| Pre-pandemic logistic sensitivity | Same fixed model; addresses contemporaneous-control misclassification | COVID: March 2020–August 2021; non-COVID: 2015–2019 | Complete demographic and PM2.5 cases | 10,887 | 12,048 |
| WUSTL PM2.5 Monte Carlo / Table 2 | Separate group-specific nulls; nominal death-relative 12–240-month means | COVID: 2020–2021; recent non-COVID: December 31, 2018–December 31, 2021 | Census-compatible age/sex and WUSTL spatial eligibility | 11,850 | 7,548 |
| AQS PM2.5 / Figure 2 | Separate group-specific nulls; death-relative 1-, 3-, and 7-year means | Same death-group date rules as the WUSTL PM2.5 analysis | AQS monitor coverage and exposure availability | 3,374 | 1,831 |
| CMAQ NO2 / Table 3 | Separate group-specific nulls; 24–168-hour means and maxima | March–July 2020 first-wave period | CMAQ coverage and complete centered randomization window | 4,522 | 1,377 |
| AQS NO2 / Figure 3 | Separate group-specific nulls; 24–168-hour monitor-based exposure | AQS input: January 2015–December 2021 | 4-km monitor coverage, exposure availability, and complete centered window | 1,206 | 659 |
| CMAQ location/time robustness | Spatial versus acute temporal decomposition | March–July 2020 first-wave period | Same eligible COVID CMAQ sample | 4,522 | — |

Notes: Counts are analysis-specific and should not be summed. The Table 2/Figure 2 recent non-COVID group is a negative-comparison group rather than the contemporaneous exchangeable comparison used by the primary logistic model. Nominal WUSTL look-backs for deaths after December 2020 use available monthly estimates through December 2020.

**Supplementary Table S2 Datasets for air pollution estimates**

| | WUSTL | EPA AQS | EPA CMAQ | EPA AQS |
|---|---|---|---|---|
| Type | Model (V5GL02) | In Situ | Model (COVID-19) | In Situ |
| Pollutant | PM2.5 | PM2.5 | NO2 | NO2 |
| Period | Jan-00 to Dec-20 | Jan-00 to Dec-21 | Jan-20 to Jul-20 | Jan-15 to Dec-21 |
| Frequency | Monthly | Quarterly | Hourly | Hourly |
| Resolution | 0.1 µg/m³ | 0.1 µg/m³ | 0.001 ppb | 0.1 ppb |
| Missing Data | 4.6% | 26.0% | 0% | 23.5% |
| Spatial Resolution | 0.01° × 0.01° | Point | 0.145° × 0.109° | Point |
| Cell Dimensions (m) | 830 x 1,110 | 4,000 (radius) | 12,040 x 12,080 | 4,000 (radius) |
| Cells in Study Area (n) | 2,867 | 11 | 32 | 2 |
| Spatial Coverage | 100% | 21.9% | 100% | 4.0% |

**Supplementary Figure S1 Study area and data coverage**

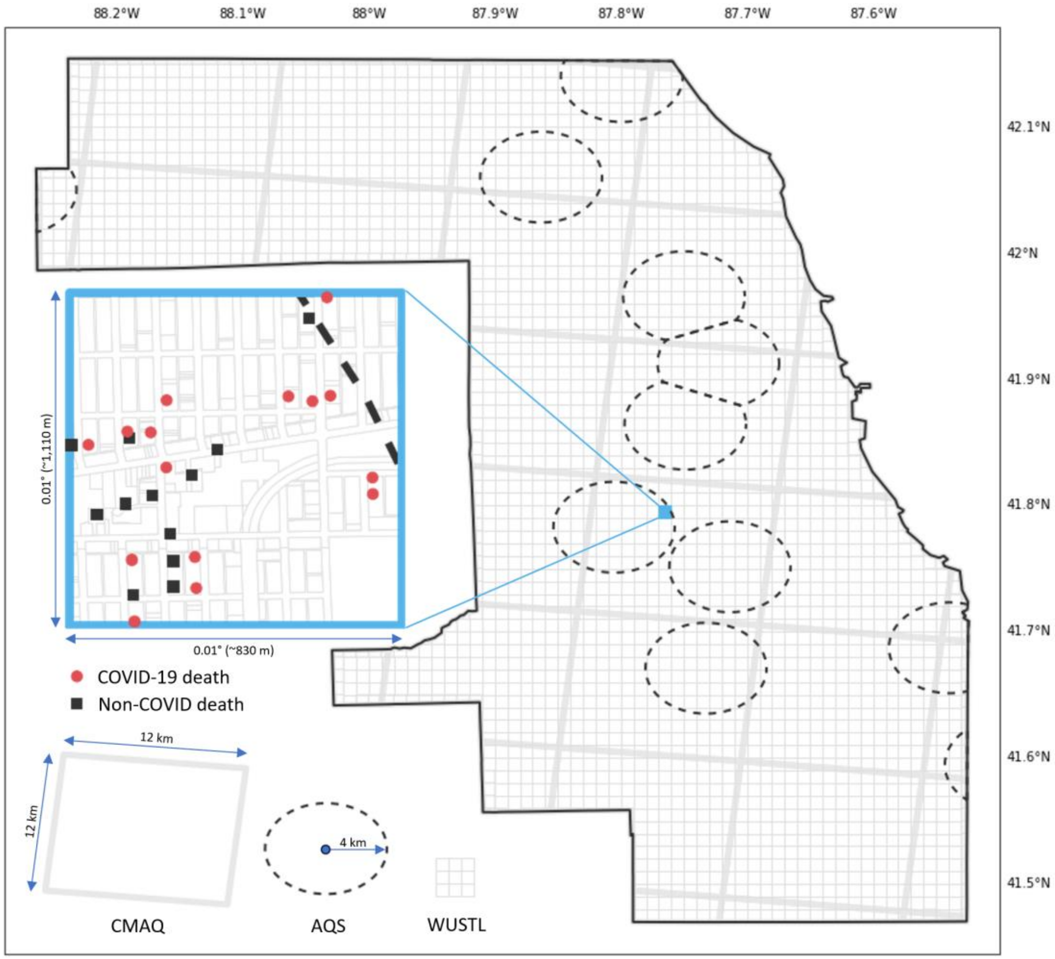


Supplementary Figure S1. Study area and data coverage in Cook County, Illinois. The inset is illustrative: symbols depict the spatial assignment of pollution estimates to COVID-19 and non-COVID death records and do not represent actual individual death locations. Dashed outlines show 4 km radius AQS monitor coverage areas; the coarser grid shows 12 km square CMAQ cells; and the finer grid shows 0.01° × 0.01° WUSTL cells.

**Supplementary Figure S2 Exposure by death category**

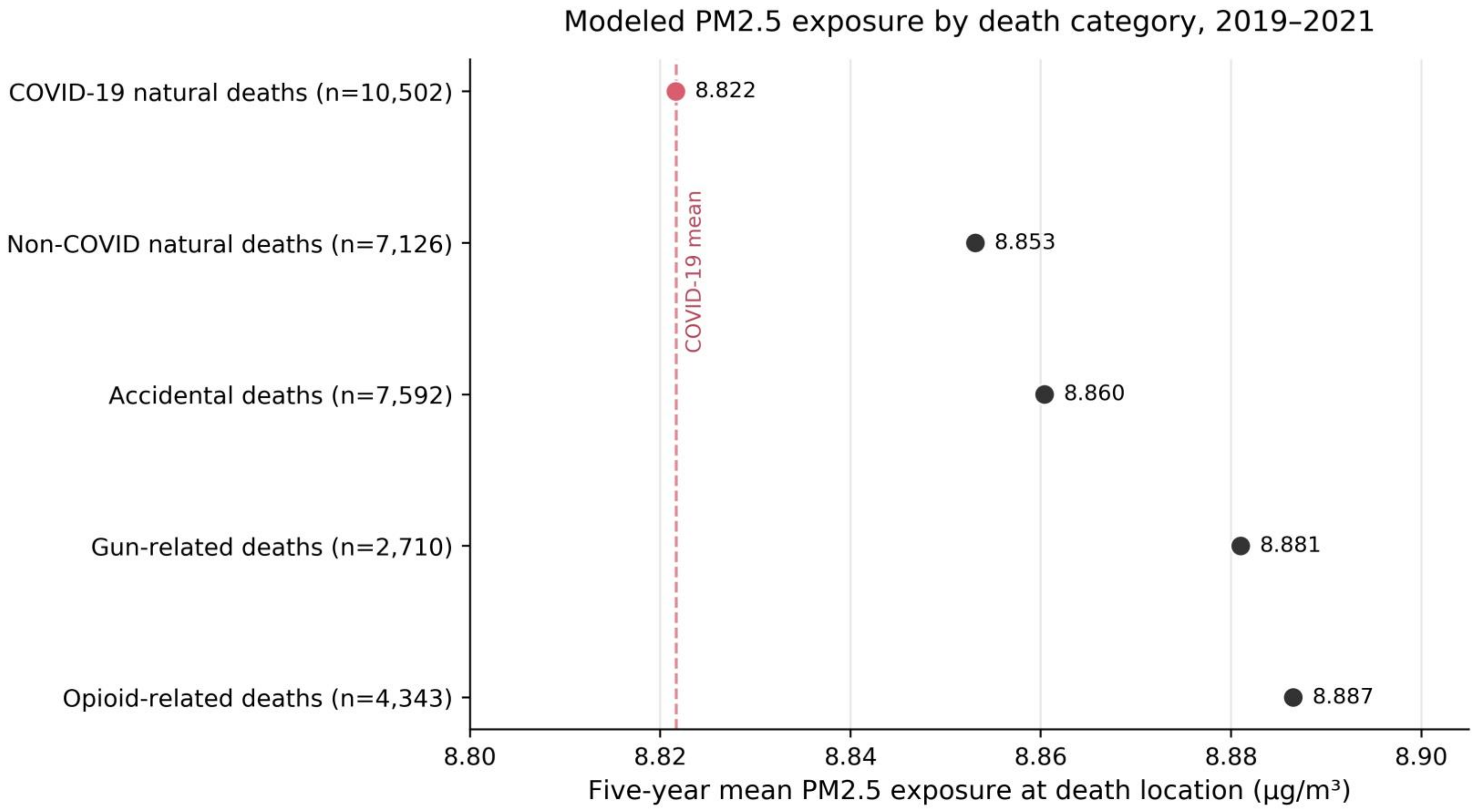


Supplementary Figure S2. Descriptive fixed five-year mean WUSTL-modeled $PM_{2.5}$ exposure ending in 2020 at Medical Examiner death locations, by death category, 2019–2021. Values are unadjusted category means; sample sizes are shown in the figure. COVID-19 and non-COVID groups are restricted to natural deaths. Gun- and opioid-related categories are flags and may overlap with accidental and other manner-of-death categories. The figure is descriptive and does not estimate causal effects or population-level mortality risk.
